# The Russian FSHD registry: a first look at the cohort

**DOI:** 10.64898/2026.03.31.26349837

**Authors:** Anna Kuchina, Darya Sherstyukova, Artem Borovikov, Margarita Soloshenko, Nikolay Zernov, Dmitrii Subbotin, Elena Dadali, Inna Sharkova, Galina Rudenskaya, Sergey Kutsev, Mikhail Skoblov, Aysylu Murtazina

## Abstract

**Background:** Facioscapulohumeral muscular dystrophy (FSHD) is a common hereditary neuromuscular disorder. The Russian FSHD Patient Registry was established in 2019 following the development of a PCR-based method for genetic confirmation of the diagnosis.

**Results:** The registry included 470 participants (51% male). Genetic confirmation was obtained for 76% (n=356), the remainder were included based on clinical and anamnestic data. Clinical assessment forms and patient-reported questionnaires were analyzed for 310 and 142 patients, respectively. D4Z4 repeat unit (RU) distribution showed patterns consistent with European cohorts, with a predominance of patients with 3 RUs. A moderate inverse correlation was found between RUs number and clinical severity scales. Periscapular weakness was the most common onset manifestation (46.8%), followed by facial weakness (31.6%) which was often unnoticed by patients. The mean age in the Russian cohort was 37.8 years (range 0–97), indicating a younger cohort compared to international data. A delta-adjusted cluster analysis (n=215) identified three distinct trajectories: a classic phenotype with onset before age 14 and early involvement of various muscle groups (n=177), and two clusters characterized by either facial or periscapular onset with slow progression.

**Conclusion:** The Russian FSHD registry provides a comprehensive characterization of a large national cohort, revealing a predominance of patients with 3 D4Z4 repeats and a younger demographic profile compared to international data. Cluster analysis identified three heterogeneous disease trajectories, offering a framework for improved patient stratification.

## 1 Introduction

Facioscapulohumeral muscular dystrophy (FSHD) is among the most prevalent hereditary myopathies. Its core clinical presentation involves progressive, often asymmetrical weakness and atrophy initiating in the facial (facio-), scapular stabilizer (scapulo-), and upper arm (humeral) muscles, with frequent subsequent involvement of the abdominal and leg muscles [1, 2]. The disease exhibits remarkably variable expressivity, with an age of onset ranging from early childhood to late adulthood and significant differences in severity even among affected family members, including instances of incomplete penetrance [3–8]. The disease has a high prevalence ranging from 2.03 to 6.8 per 100,000 individuals in different countries [9, 10].

Substantial progress in elucidating the pathogenetic mechanisms underlying FSHD, most notably the discovery of epigenetic derepression of the D4Z4 array and the subsequent toxic gain-of-function of DUX4 expression, has paved the way for the development of novel targeted therapies [11–13]. The advancement of these promising interventions underscores the necessity of well-established patient registries. Such registries are crucial for standardizing data collection, enabling efficient access to consolidated clinical information, and supporting natural history investigations and the planning of clinical trials in real-world contexts. At present, more than 20 FSHD registries operate globally [14].

For an extended period, a national FSHD registry had not been established in Russia, largely due to unavailability of molecular diagnostics. In the absence of accessible molecular testing, diagnoses were predominantly based on clinical presentation and family history. Southern blotting, the standard method for quantifying D4Z4 repeat units (RUs) [15–17], is not used in routine Russian diagnostic practice due to restrictions on the use of radioactive isotopes. To address this diagnostic gap, a PCR-based method was developed and locally validated, and it has been successfully employed for the molecular confirmation of FSHD since its publication in 2019 [18]. This advancement subsequently facilitated the establishment of a national FSHD patient registry at the Research Centre for Medical Genetics (RCMG) (https://fshd.med-gen.ru/en/).

In this article, we present the first description of the Russian registry data as of January 2026. We provide a comprehensive description of the registry cohort, including clinical and genetic characteristics, and statistical analysis of the collected data.

## 2 Methods

### 2.1 Platform and data collection

All participants of the Russian FSHD patient registry or their legal guardians provided informed consent before inclusion in the registry. The registry is based on the specialized software platform, used to automate the work of medical organizations and create a unified information space for doctors with separate access to data for each employee. A schematic diagram illustrating the formation of the register is provided in **Fig. 1a**.

**Fig. 1.**
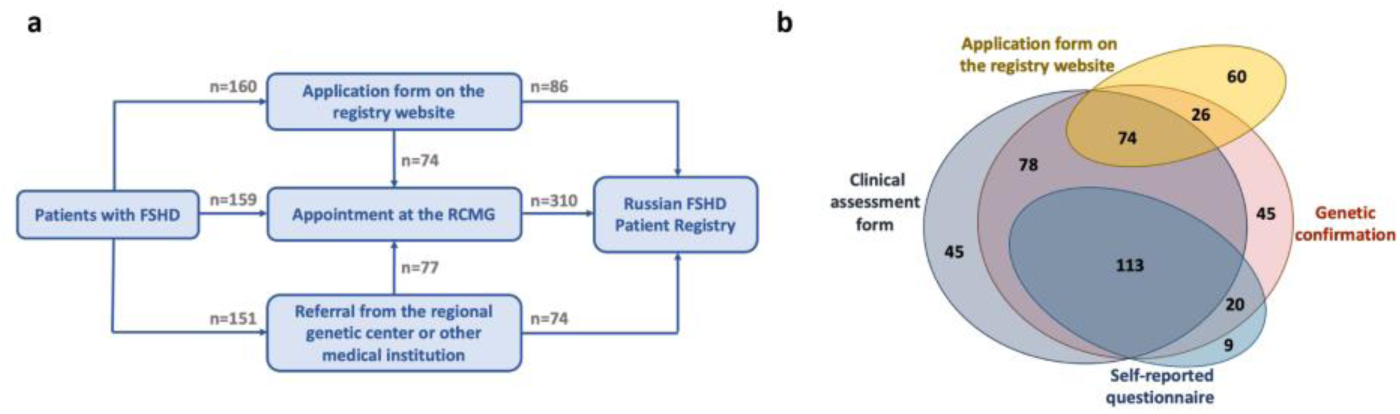
Registry flowchart and structure. (a) The pathways for including patients in the FSHD registry. (b) Venn diagram illustrating the distribution of registry participants based on the availability of clinical assessment form, patients self-reported questionnaire and genetic confirmation

Patients were included in the FSHD registry through three main pathways:

1. ***Referral:*** A patient was referred by a regional genetic center or other medical institution.
2. ***Self-Application:*** The patient completed an application form on the registry website, which contained specially designed questions to identify clinical signs indicative of FSHD. Patients with symptoms characteristic of FSHD were identified. They were asked to visit the center for a more precise diagnosis or, if unable, to provide additional clinical and genetic information about their condition.
3. ***Patients referred for genetic consultation to the RCMG:*** Patients referred with a diagnosis of unspecified myopathy were identified by a geneticist during a genetic consultation at the RCMG and subsequently registered.

Thus, the registry includes patients with a genetically confirmed diagnosis, as well as patients without genetic confirmation, but with a typical clinical presentation of FSHD.

Clinical data was gathered via two primary sources: a patient-reported questionnaire and a detailed clinical examination performed during patient’s visits **(Fig. 1b**). Twenty nine patients completed a patient-reported questionnaire without detailed clinical examination at our center, 197 patients were examined without filling out the self reported questionnaire, and 113 patients passed both assessments.

### 2.2 Clinical assessment form

We developed a specialized, 196-item clinical assessment form for standardized patient data collection. The form is structured into seven comprehensive sections: (I) basic patient information; (II) genetic diagnostic results; (III) anamnestic data (IV) neurological examination; (V) assessment via specialized clinical scales; (VI) extramuscular signs; (VII) laboratory and instrumental studies. The contents of the sections of the Clinical assessment form are presented in the Supplementary table 1. For clinical assessment the FSHD clinical score (FSHD-CS) and the 10-grade Сlinical severity scale (CSS) were applied, as well as age corrected FSHD-CS and CSS as it was previously suggested [19–21]. Moreover, patients were classified according to the Сlinical categories used in the FSHD Comprehensive Clinical Evaluation Form [22, 23].

### 2.3 Patient-reported data

We developed a 62-item questionnaire for self-assessment of a disease course. The items encompassed patient-reported symptoms, complaints, and factors influencing disease progression. Two self-reported scales to assess patients’ quality of life, the Facial Disability Index (FDI) and *GNE*-myopathy functional activity scale (GNEM-FAS) were added to the self-reported questionnaire [24, 25]. The latter was originally developed to evaluate functional ability (mobility, upper limb use, and self-care) in adults with *GNE*-myopathy. Although initially developed for *GNE*-myopathy, we found this scale well-suited for assessing functional status in patients with other myopathies, including FSHD.

### 2.4 Genetic diagnostics

For patients with a genetically confirmed diagnosis, confirmatory testing was performed either at the RCMG or at specialized international genetics centers, with results provided for documentation. Genetic analysis was conducted on DNA extracted from peripheral blood. In five cases, cultured skin fibroblasts were used for diagnostic confirmation. All assays employed the previously developed and described methodology [18], enhanced with recent qualitative improvements. Due to the specificities of the assay methodology, there are instances where the precise number of RUs cannot be determined. In such cases, the result is reported as a range between adjacent numbers (e.g., 3-4 or 5-6).

In brief, the methodology comprises four main steps:

1. Isolation of high-molecular-weight (HMW) DNA from PBMCs or fibroblasts in agarose plugs, as previously described [18]. Now, only 4 mL of fresh, unfrozen blood is required to obtain 10–20 plugs (approximately 1–2×10^6 cells per plug), or at least 5×10^5 fibroblast cells per plug.
2. Restriction enzyme digestion of DNA using EcoRI to excise D4Z4 arrays originating from chromosomes 4 and 10.
3. Pulse-field gel electrophoresis (PFGE), followed by gel staining and fragmentation to isolate DNA segments of specific lengths corresponding to particular repeat numbers of D4Z4. The PFGE conditions were slightly modified compared to the original protocol and are as follows: 2.5 L of 0.5× TBE buffer; voltage 450 V; pulse time 0.5 sec; total duration 2 hours 45 minutes; buffer temperature +10 °C. The molecular weight markers used for fragment sizing were the Lambda Mix Marker (Thermo Scientific, Waltham, MA, USA) and the M12 DNA Ladder (SibEnzyme, Novosibirsk, Russia). The gel fragmentation scheme was revised and is provided in the Supplementary **Fig. S1**.
4. Multiplex quantitative PCR (qPCR) with two pairs of specifically designed primers for detecting both the number of D4Z4 repeats on chromosome 4 alleles and the permissive haplotype A of these repeats. A total of 10 µL of each of the fivefold-diluted gel fragments after PFGE served as templates. Negative and positive PCR controls followed the previously established protocols. PCR conditions included an initial “hot-start” step at 95 °C for 3 minutes, followed by 40 cycles of: 95 °C for 30 seconds, 62 °C for 5 seconds, and 72 °C for 15 seconds. Reactions were performed in a final volume of 25 µL containing: 1.25 U of “SmarNGTaq” DNA polymerase (Dialat Ltd., Moscow, Russia); 1× “Ampli” PCR buffer (Dialat Ltd., Moscow, Russia); 1.2 M Betaine (Tokyo Chemical Industry Co., Ltd., Tokyo, Japan); 5 mM MgCl₂ (Dialat Ltd., Moscow, Russia); and dNTPs (0.2 mM each of dATP, dTTP, dCTP, and dGTP). Primers and TaqMan probes were used at equimolar concentrations, 10 picomoles each per reaction. The primers for detecting chromosome 4q D4Z4 repeats (RU) were as follows: Rep1,5F: 5′-GTGCTTGCGCCACCCACGT-3′; Rep1R: 5′-GCCGCGCGGAGGCGGAG-3′; TaqMan probe (zond2Rep1): 5′ FAM-AGTCCGTGGTGGGGCTGGGG-BHQ1 3′. Primers for detecting the permissive haplotype A of 4qD4Z4 were: AS_pLAM: 5′-CACAGGGAGGGGGCATTTTA-3′; pLAM-FW: 5′-TCTGTGCCCTTGTTCTTCCGT-3′; TaqMan probe (ZondpLAM3.2+): 5′ Rox-TGGCTGAATGTCTCCCCCCACCT-BHQ2 3′ qPCR was performed in duplicate for each gel fragment. Data analysis employed the 2^−ΔΔCt method.

### 2.5 Statistical analysis

Statistical analysis was performed using Python 3.12 with the pandas, scipy, and statsmodels libraries. Descriptive statistics were reported as medians with interquartile ranges for non-normally distributed continuous variables and as frequencies and percentages for categorical data. Correlations between D4Z4 repeats and clinical indices (age-corrected FSHD-CS and age-corrected CCS) were assessed using Spearman’s rank correlation. Categorical associations were evaluated via Chi-square or Fisher’s exact tests, with effect sizes reported as Cramér’s V. Multiple testing was corrected using the Benjamini-Hochberg procedure.

To characterize patterns of symptom onset progression, hierarchical clustering was applied to 215 patients with at least one symptom of known onset age. For each patient and each of five anatomical groups (facial, periscapular and shoulder, peroneal, pelvic girdle and thigh, and axial), a delta value was computed as the difference in years between the onset age of that symptom group and the patient’s earliest recorded symptom onset. Missing delta values, present in 12–83 patients per group depending on anatomical region, were imputed using an iterative imputer (MICE-analogous multivariate imputation, 20 iterations, median initialization) incorporating age at examination and first symptom age as auxiliary predictors. Ward’s minimum variance method with Euclidean distance was used for agglomerative hierarchical clustering of the resulting 215 × 5 imputed delta matrix. The optimal number of clusters was evaluated using the average silhouette coefficient, three clusters were retained to provide clinically interpretable subgroup granularity (silhouette = 0.493 for k = 3).

## 3 Results

### 3.1 General information

As of January 2026, the FSHD patient registry contained data from 470 individuals across 388 families (388 probands and 82 affected relatives). Of these patients, FSHD1 was confirmed in 346 patients from 276 families, and FSHD2 in 10 patients from 8 families. For 114 individuals from 104 families, enrollment in the registry was based on clinical examination alone, without genetic confirmation at the time of registration. The gender distribution was nearly equal (241 male, 51%). The median age of the patients was 38 years, with a range spanning from 0 to 97 years. Patients were most frequently represented in the 18–40-year (n=205) and >40-year (n=206) age groups (**Fig. 2a**). The pediatric group (age <18 years) was smaller (n=59; 12.5%).

**Fig. 2.**
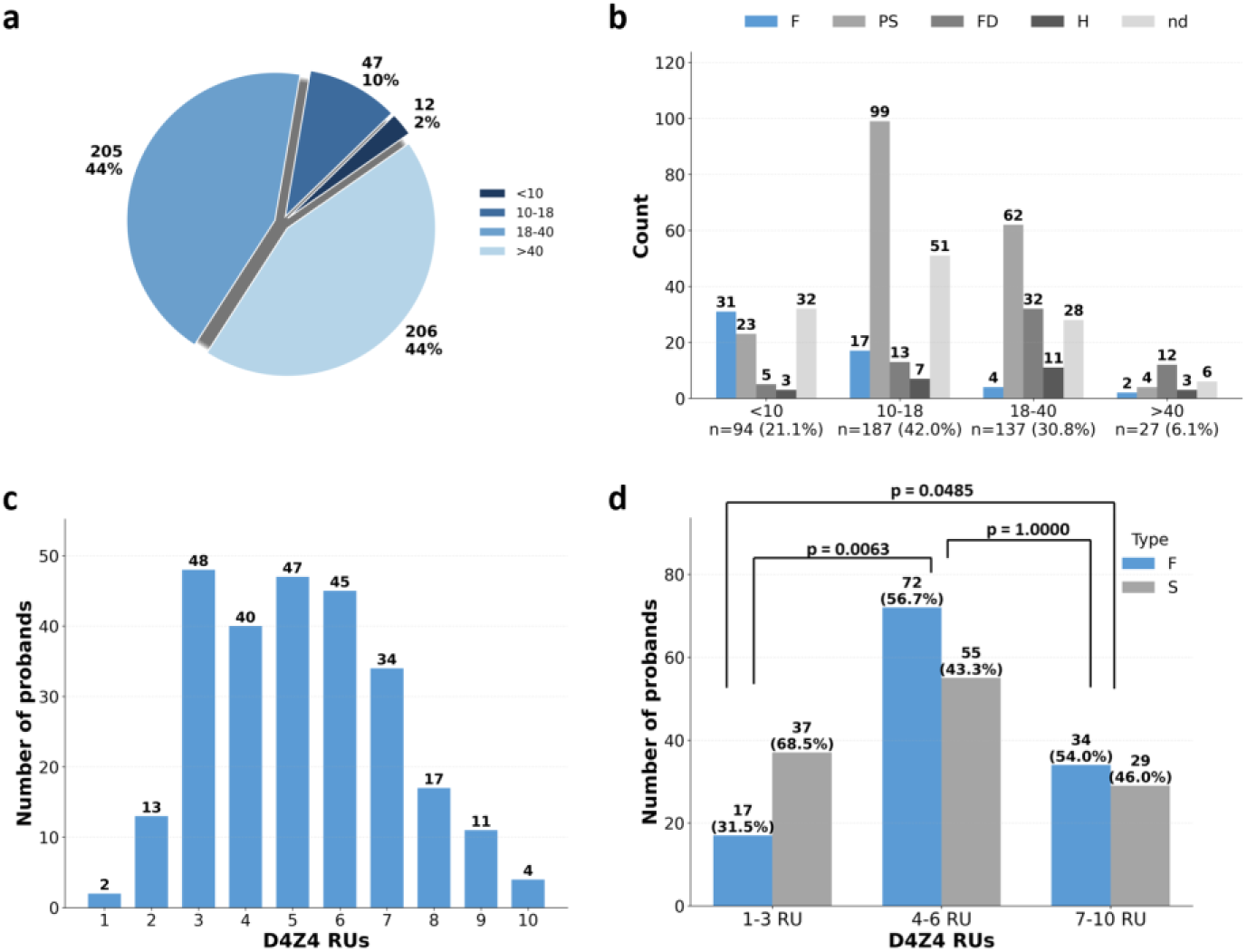
General characteristics of the Russian FSHD patient registry. (a) Patients’ age distribution, presented as count (n) and percentage (%). (b) Distribution of patient-reported disease onset age and initial symptom location (n=445). (c) The distribution of D4Z4 RUs in probands. Due to the limitation of genetic analysis, in some cases the results were reported as a range between adjacent numbers (e.g., 3-4 or 5-6). For the purpose of graphical representation and analysis, these indeterminate cases were excluded. The figure displays data only from families with a precisely determined RUs number (n=261). (d) The distribution of sporadic and family cases in the D4Z4 range groups. RUs - repeat units, F - facial, PS - periscapular, FD - foot dorsiflexion, H - hip, nd - no data

A family history analysis revealed that 259 cases had affected relatives, 167 were sporadic, and in 44 cases, the family history was unclear. The ambiguity in these cases was due to incomplete pedigree data (e.g., single-parent families, early parental death) or uncertainty regarding the presence and nature of symptoms in relatives based on patient report, which precluded a definitive FSHD diagnosis.

Patient-reported age of onset and initial symptom localization were classified into groups, with the distribution shown in **Fig. 2b**. This analysis was performed on 445 patients. A total of 25 patients were excluded due to unclear age at onset (n = 18) or presymptomatic status (n = 7). The largest group comprised patients with onset between 10 and 18 years (n=187). Among these, weakness of the periscapular muscles was the most common initial symptom (n=99). A notable proportion had difficulty localizing the first symptom (n=51). Less frequent presentations included facial muscle weakness (n=17), foot dorsiflexion weakness (n=13), and proximal lower limb weakness (n=7).

The second largest group had symptom onset between 18 and 40 years (n=137). In this group, periscapular weakness was again the most prevalent initial symptom (n=62). However, foot dorsiflexion weakness at onset was more common here (n=32) than in the younger (10-18 years) group. Difficulty localizing the first symptom was reported by 28 patients, while proximal leg weakness (n=11) and facial weakness (n=4) were less frequent.

The third group included patients with onset before age 10 (n=94). This cohort more frequently reported facial muscle weakness at onset (n=31), along with difficulty localizing the first symptom (n=32). Periscapular weakness was less common (n=23), and initial symptoms affecting foot dorsiflexion (n=5) or the proximal lower limb (n=3) were rare.

The smallest group consisted of 27 patients with onset after 40 years. Here, foot dorsiflexion weakness was the most frequent initial symptom (n=12), followed by: difficulty localizing the first symptom (n=6), periscapular weakness (n=4), hip weakness (n=3), and facial weakness (n=2).

Finally, the cohort included seven asymptomatic individuals with a D4Z4 contraction identified through familial screening.

Seventy-one patients (15% of the cohort) presented with limited mobility. Among them, 29 patients used mobility aids (e.g., canes, walkers, or orthoses). A total of 42 patients (59.2% of this subgroup) were partially or fully wheelchair-dependent, including three who were under 18 years of age.

### 3.2 Genetic studies results

Genetic studies were performed for the majority of patients, comprising 356 registry participants (75.7%). Among these, FSHD1 was diagnosed in 346 individuals from 276 families. The distribution of D4Z4 RUs in these probands is shown in **Fig. 2c**.

A comparative analysis of familial and sporadic cases among probands with FSHD1 **(Fig. 2d)** revealed statistically significant differences (χ²=10.064, p=0.00653) showing the predominance of sporadic cases in the group of patients with 1–3 RUs compared to the groups with 4–6 RUs and 7–10 RUs (p-value = 0.00626 and p-value = 0.04850, respectively). No significant differences were found between the groups with 4–6 RUs and 7–10 RUs (p-value(Bonferroni) = 1.00000).

Detailed data on patients with confirmed FSHD2 will be reported in a subsequent manuscript (currently in preparation).

### 3.4 Clinical data

#### 3.4.1 Patient self-reported data

A total of 79 male and 63 female patients or their legal guardians completed the questionnaire. At the time of data collection, the mean patient age was 32.8 years (range: 5–92), and the mean patient-reported age at disease onset was 17.7 years (range: 0–65).

The assessment of comorbid status revealed a wide spectrum of comorbid diseases or conditions among 49 patients (34.5%). Gastrointestinal disorders were the most frequent, identified in 18 cases (12.7%), including chronic gastritis (n=8), pancreatitis (n=3), chronic cholecystitis (n=3), a history of hepatitis A or C (n=3), and irritable bowel syndrome (n=1).

Endocrine disorders were present in 14 patients, primarily thyroid pathologies (n=9), among which autoimmune thyroiditis was noted. Less common were diabetes mellitus and insulin resistance. Cardiovascular conditions were identified in 9 cases, with arterial hypertension predominating (6 cases). Isolated instances of congenital heart disease, cardiomyopathy, and arrhythmias were also recorded.

Oncological conditions, both benign and malignant, were reported in 4 cases, including pituitary microadenoma, mammary fibroadenoma, and papillary thyroid carcinoma. Dermatological and allergic diseases were relatively uncommon (4 cases), such as allergic dermatitis, bronchial asthma, and allergic rhinitis. Finally, isolated cases of systemic lupus erythematosus and chronic renal failure were also noted.

The survey revealed that approximately one-third of patients (n=49; 34.5%) were involved in some form of sport or physical activity at the time of data collection. The majority preferred low-impact activities, such as therapeutic exercise, swimming, fitness, yoga, or gym training (n=39). However, some participants reported more intense or high-risk sports, including tennis, volleyball, choreography, snowboarding, martial arts, and motocross. When asked about physical activity prior to disease onset, the majority of respondents (n=87; 61.7%) reported having been involved in sports, including at a professional level. Nevertheless, half of the patients (n=73) indicated experiencing significant difficulties in performing physical exercises during their school education.

Approximately one-third of participants (35.2%, n=50) associated the development of the disease with a specific trigger. For example, 12 patients identified intense physical exercise as a trigger, while one patient attributed the cessation of physical exercise to the onset of the disease. Of the 30 women whose medical records indicated pregnancy, half reported a deterioration in their general condition during that time. Furthermore, pregnancy itself was identified as a precipitating factor for disease onset in four patients, and childbirth in two others. Other reported triggers included emotional stress (n=11), surgery (n=7), trauma (n=7), hormonal changes (n=5), and weight loss (n=1).

At disease onset, 98 patients (69%) reported asymmetrical involvement, with a predominance of right-sided weakness (ride side: n=69; left side: n=29). By the time of the survey, asymmetry was reported by a larger proportion of patients (n=111; 78.2%; right side: n=76; left side: n=35). A small proportion of patients (n=35; 24.6%) reported experiencing pain of varying intensity in different parts of the body prior to the onset of muscle weakness.

The mean score on the physical subscale of the Facial Disability Index (FDI) was 90.67 (SD=±12.73, range: 45–100). On the GNEM-FAS, the mean total score was 79.27 (SD=±19.42, range: 19–100), with subscale means of 28.71 for Mobility (SD=±10.45, range: 1–40), 27.19 for Upper Extremity (SD=±5.11, range: 7–32), and 23.37 for Self-Care (SD=±5.21, range: 5–28).

#### 3.4.2 Clinical assessment form

Clinical data via clinical assessment form were collected from 310 patients, comprising 265 confirmed cases and 45 cases without molecular genetic confirmation **(Fig. 1b)**. The mean age at examination was 33.9 years (range: 5–83 years), with 61 patients (19.7%) being under 18 years of age. The cohort included 260 probands and 50 their relatives.

The mean age of onset was 15.39 years (SD=±12.36). Regarding symptom localization at onset, patients most frequently self-reported weakness of the periscapular muscles (n=170; 54.8%). This was followed by weakness in the lower leg (n=59; 19.0%) and facial weakness (n=46; 14.8%). Onset of weakness of the thigh and pelvic girdle muscles was observed in 21 patients (6.8%). A smaller group (n=11; 3.5%) had difficulty naming the location or were presymptomatic. Isolated cases with onset in the abdominal (n=2) or neck muscles (n=1) were also noted. Following careful collection of anamnestic data, the distribution of initial symptom localization shifted. Weakness of the periscapular muscles remained the most common presentation, though less frequent (n=145; 46.8%). Facial muscle weakness emerged as the second most common localization (n=98; 31.6%), while lower leg weakness moved to third place (n=39; 12.6%). Onset in the pelvic girdle was identified in 12 patients (3.9%), and with weakness of the abdominal muscles in 9 patients (2.9%). The remaining 7 patients included presymptomatic individuals, those with indeterminate symptom onset, and one patient with neck muscle weakness. Asymmetrical involvement, manifesting as unilateral muscle atrophy, facial asymmetry, or asymmetric scapular winging, was observed in 84.5% of cases.

Muscle weakness among patients was distributed as follows. Most exhibited weakness of the facial muscles, primarily the orbicularis oris and cheek muscles (n=283; 91.3%). Weakness of the orbicularis oculi muscle was present in 78.4% of cases (n=243). Pectoral muscle weakness occurred in approximately half of the patients (n=161; 51.9%). Neck muscle weakness was relatively uncommon. Weakness of the neck flexors was observed only in one-tenth of patients (10.3%, n=32) and was predominantly mild to moderate (MRC grade 3–4/5, n=28); four cases showed severe weakness of neck flexors (MRC grade 0–1/5). Among patients with neck flexor weakness, the mean age at disease onset was 8.8 years (range 0–41) and the mean age at examination was 29.6 years (range 5–70). In the subgroup with severe neck flexors weakness, the age of onset was much earlier (mean 3.7 years, range 0–10). Weakness of the neck extensors was even less frequent, occurring in only 2.3% of patients (n=7), and was also mild to moderate in severity.

In the shoulder girdle, the most commonly involved muscles were the supraspinatus (56.8%) and infraspinatus (57.7%) muscles followed by the biceps (36.1%) and triceps (21.9%). The deltoid muscle was intact in most cases (87.1%), as well as the hand fingers extensors and wrist extensors (83.2% and 88.4%, respectively). Involvement of the hand fingers and wrist flexors was rare (<5%).

The primary pattern of involvement in the pelvic girdle and lower limbs according to clinical assessment highlighted the following muscle groups: the tibialis anterior muscle (51.3%), the hip extensors (45.5%), and the foot muscles (extensor hallucis brevis - 46.1%, extensor digitorum brevis - 41.0%). The remaining muscle groups were involved substantially less frequently. Muscles such as the quadriceps femoris, adductors, abductors, and triceps surae were affected significantly less often (in less than 23% of cases).

Two-dimensional principal component analysis (PCA) was performed on all patients with a clinical assessment utilizing manual muscle testing scores for the following muscle groups: facial, shoulder girdle, pelvic girdle and thigh, and lower leg muscles **(Supplementary, Fig. S2)**. No clear distribution was observed based on either the age of disease onset or disease duration on obtained PCA pattern.

Scapular winging was identified in 292 patients (94.1%). Beevor’s sign, indicative of abdominal muscle weakness, was present in 68.4% of cases (n=212). Notably, while upward umbilicus displacement was observed in most of these (n=198, 93.4%), downward displacement was recorded in 14 cases (6.6%).

Camptocormia was an uncommon finding, present in only five patients. In one of these cases, family history indicated relatives with FSHD who also exhibited camptocormia. The mean age of this subgroup was 57.8 ±17.8 years (range 30–74), with D4Z4 repeat contractions ranging from 6 to 8 units. The mean disease onset age within this group was 24 ±8.2 years (range 16–25). All patients with camptocormia presented with scapular winging, facial muscle involvement, and a positive Beevor’s sign; none exhibited neck flexor or extensor weakness.

Extramuscular manifestations included sensorineural hearing loss of varying severity (1 of 18 patients who provided a specific examination). However, upon clinical examination, 21 patients complained of hearing loss and were referred to otorhinolaryngologist. Retinal angiopathy was detected in 10 of 50 patients who provided this examination (19.2%). Cataracts were detected in 6 cases and isolated cases of retinal detachment and partial optic nerve atrophy. Spirometry data were obtained from 14 patients. Pulmonary ventilation impairments were detected in 8 of them (57%). Five patients had a mild to moderate decrease in vital capacity. Three patients showed moderate restrictive impairment. Among 108 patients who provided electrocardiography or echocardiography results, changes were identified in 17 cases. Cardiac rhythm or conduction abnormalities were most commonly identified, including sinus bradycardia, sinus bradyarrhythmia, sinus arrhythmia, supraventricular extrasystole, atrial fibrillation, incomplete right bundle branch block, and Wolff-Parkinson-White syndrome. In single cases, cardiovascular diseases with structural myocardial involvement were diagnosed, namely ischemic heart disease and arterial hypertension accompanied by left ventricular myocardial hypertrophy and left atrial dilatation. Single cases of ventricular septal defect and atrial septal aneurysm were also observed. Detailed clinical characteristics of patients are presented in the Supplementary table 2.

Correlation analysis between D4Z4 RUs number and disease severity scores revealed a moderate inverse association between RUs number and age-corrected FSHD-CS score with high statistical significance (Spearman correlation coefficient ρ=-0.546, p-value=1.89×10⁻²¹), as well a moderate inverse association between RUs number and age-corrected CSS score (Spearman correlation coefficient ρ=-0.531, p-value=3.91×10⁻²⁰ **(Fig. 3a)**.

**Fig. 3.**
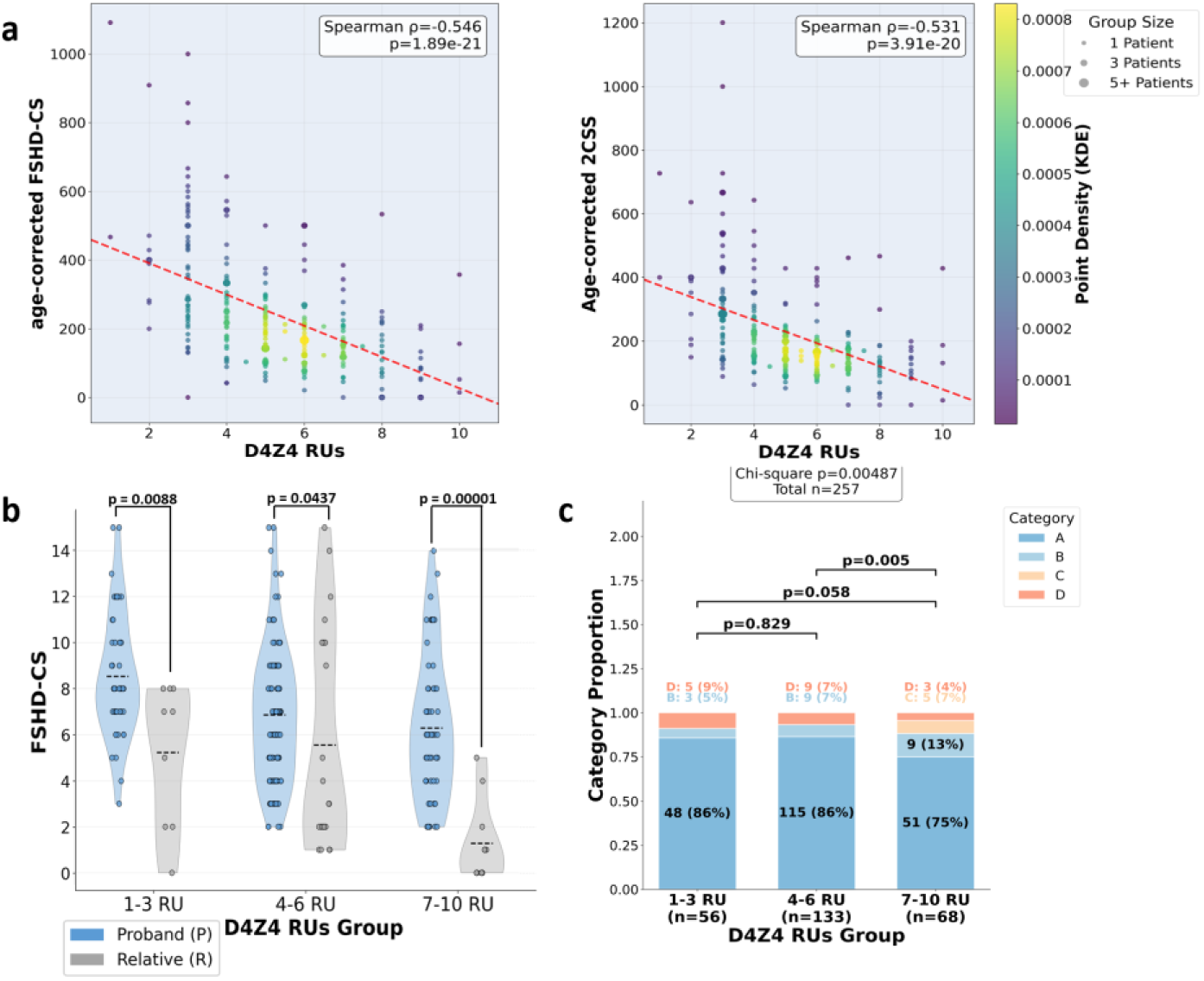
Clinical and genetic characteristics of patients with FSHD1. (a) Correlation analysis between numbers of D4Z4 RUs and clinical severity scales. (b) Comparison of disease severity using the FSHD-CS between probands and their relatives, stratified according to D4Z4 RUs. Subjects were subdivided by D4Z4 RUs: 1–3, 4–6, 7–10 RUs. (c) Distribution of cases according to D4Z4 RUs and clinical categories. In each subgroup, the proportion of cases who were assessed as clinical category A, B, C or D are reported

The comparison between clinical severity of FSHD in probands and their relatives, stratified by D4Z4 RUs range, are shown in **Fig. 3b**. A statistically significant difference in clinical severity was found between probands and their relatives in all groups analyzed, with probands consistently presenting with a more severe phenotype. In the 1-3 RUs group, the mean FSHD-CS score was higher for probands (mean = 8.5 ± 2.63, range 3–15) compared to their relatives (mean = 5.2 ± 3.11, range 0–8). In the 4-6 RUs group, the mean scores were more similar between probands and relatives (6.8 ± 2.87 and 5.5 ± 4.48, respectively). In the 7-10 RUs group, a significant difference was observed, with probands having a higher mean FSHD-CS score (6.3 ± 3.12) compared to their relatives (1.3 ± 3.22).

The distribution of patients across clinical categories (A-D) for each RUs range is shown in **Fig. 3c**. Category A (typical FSHD phenotype) was predominant across all groups, comprising 86% (n=48) of the 1-3 RU group, 86% (n=115) of the 4-6 RU group, and 75% (n=51) of the 7-10 RU group. Category B (incomplete FSHD phenotype) was less represented: 5% (n=3), 7% (n=9), and 13% (n=9) of the respective groups. Category D (atypical signs) was present in 9% (n=5), 7% (n=9), and 4% (n=3) of cases, respectively. Notably, Category C (subjects without motor impairment or with minor possibly FSHD-linked signs) was observed only in the 7-10 RU group. A statistical difference in the distribution of clinical categories was revealed when comparing the groups of 4-6 RUs and 7-10 RUs (p-value = 0.005).

#### 3.4.3 Clustering of patients by weakness involvement pattern

To identify patient subgroups, clustering was performed based on the sequence of symptom progression, focusing on the order in which different muscle groups were affected. These included facial, periscapular and shoulder, axial, pelvic girdle and thigh, and peroneal muscle weakness. To account for individual differences in the timing of symptom onset, the delta score was included in the model as an adjusting variable **(Fig. 4)**.

**Fig. 4.**
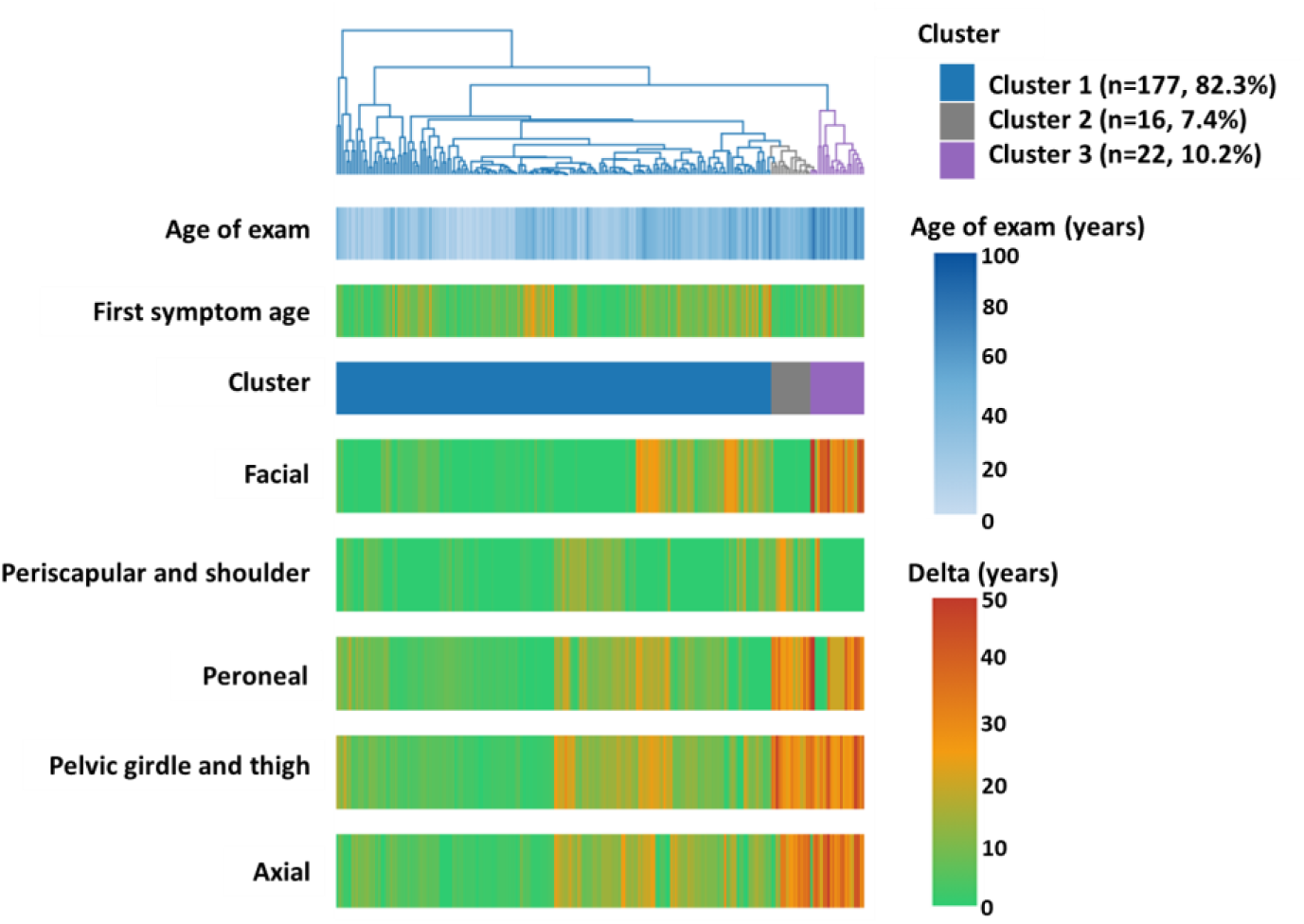
Hierarchical clustering of registry participants based on the sequence of symptom progression, focusing on the order in which different muscle groups were affected with delta score adjustment

Based on the clustering results, three clusters were identified. The largest patient group was defined by three key features (n=177): symptom onset before the age of 14.0 (IQR 6.0–21.0), a rapidly progressive disease course, and the early spread of weakness to other muscle groups (facial delta 2.0 (0.0–9.0) years, periscapular and shoulder delta 1.0 (0.0–6.0) years). This cluster was characterized by the weakness of all five muscle groups by the age of 30.

The second cluster included patients with early involvement of the facial muscles followed by periscapular muscles, but with slow progression of the weakness in other muscle groups including axial muscles, pelvic girdle and lower limb muscles (n=16, 7.4%; peroneal delta 27.5 (24.8–33.0) years, pelvic girdle and thigh delta 30.0 (26.2–34.2) years; p-value <0.001 vs Cluster 1 for other groups). The axial, pelvic girdle and thigh, and peroneal muscles delta did not differ significantly from those in Cluster 3 (p-value = 0.45, p-value = 0.78, and p-value = 0.064, respectively).

The third cluster (n=22, 10.2%) comprised cases with slow progression, primarily defined by early involvement of the shoulder girdle muscles and, to some extent, the peroneal group, with notably late involvement of all other muscle groups, including the facial muscles (facial delta 31.5 (28.2–41.0) years; p-value <0.001 vs Clusters 1 and 2).

## 4 Discussion

For 2022, the global FSHD registry network collected data on 21 national or regional registers [14]. The largest registry is the Italian national registry, which enrolled 2,169 patients as of August 2021 [26]. This constituted 26% of all registered patients worldwide. The next largest registries were UK FSHD Patients Registry (1,035 patients), Chinese Clinical Trial Registry (998 patients), the National Registry of Myotonic Dystrophy and FSHD Patients and Family Members in the USA (975 patients) and the French National Registry of Facioscapulohumeral Dystrophy (930 patients) [26]. The Russian FSHD registry, with 470 patients as of January 2026, is comparable in size to other registries and contributes to the global study of the disease. While the number of patients in our registry is significantly lower than the expected prevalence in Russia, we anticipate steady growth in the coming years.

In some registries, molecular genetic diagnostics are performed for all patients (e.g., the Italian National Registry, the Chinese Clinical Trial Registry, and the FSHD Registry in the Netherlands), whereas in others, genetic testing is not performed on all patients [26]. For example, as of 2021, the proportion of genetically confirmed cases was 96% in the French National Registry of Facioscapulohumeral Dystrophy and 50% in the UK FSHD Patient Registry. In the Russian registry, the majority of patients had genetic confirmation (75.7%); however, the remaining patients did not undergo molecular genetic testing due to an inability to reach the center for examination, challenges in providing biomaterials, or financial limitations.

Recent studies have shown that in East Asian countries, shorter FSHD1 alleles (1-6 RUs) are significantly more frequently detected among genetically confirmed patients with FSHD1 than alleles with 7-10 RUs, in contrast to the European countries [27, 28]. In our registry, the overall distribution of RUs was similar to that observed in European registries. Groups with extreme values (1–3 and 7–10 RUs) were represented by fewer patients, with the lowest frequencies observed for 1 and 10 RUs, while the 4–6 RU group was the most numerous. However, the representation of patients with 8–10 RUs in the Russian registry was lower than in European cohorts. Notably, unlike in European countries, patients with 3 RUs formed the largest group in our registry, making the distribution somewhat more similar to that seen in Asian countries [27–31].

In our study, eight families with FSHD2 were identified, comprising 2.8% of the total cohort. This proportion aligns with findings in European registries, where FSHD2 accounts for 3–6% of all FSHD cases, whereas in India, this figure reaches 8% [27, 32–34].

Our data differ from those obtained from the UK registry, where facial muscle weakness was the predominant initial symptom (52.08% vs. 31.6%), followed by periscapular muscle weakness (34.5% vs. 46.8%) [35]. However, our results correspond with those from the North Indian registry: in both groups, proximal arm weakness was the most common initial presentation (46/101; 45.5%), while facial muscle weakness, though second in prevalence, was noted in only a minority of cases (10/101; 9.9%) [27]. The discrepancy between the first symptom reported by patients and the first symptom identified after the detailed history collection highlights the importance of thorough history taking in FSHD. It also suggests that facial muscle weakness frequently remains unnoticed by patients, as it does not substantially interfere with daily activities.

In the literature, two forms of the disease with onset after 18 years of age are typically distinguished: adult-onset and late-onset. Nevertheless, no consensus exists regarding the age cutoff separating these forms, with some authors using 30 years and others using 45 years [36, 37]. In the present study, 40 years was chosen as the discriminating threshold. Overall, shoulder girdle involvement was the predominant initial symptom in the adult-onset group (onset 18-40 years), whereas peroneal weakness predominated in the late-onset group (onset >40 years), which is consistent with recent reports [37]. As previously reported, there were no late-onset (after 45 years) patients with initial facial weakness in our cohort [37].

Notably, the proportion of the patients with limited mobility in the Russian registry was less than in other registries. For example, the proportion of patients using walking aids in the Russian registry was 6.2%, compared with 36.9% and 29% in the Dutch and UK registries, respectively [38, 39]. The proportion of individuals using a wheelchair part-time or full-time for mobility also varied: in our registry, it was 6.3% and 3.9%, respectively, compared to 22.2% and 17.7% in the UK registry of the entire group of patients with FSHD. In the Dutch registry, the corresponding proportions were 28% and 11%, respectively. The disability index can be influenced by various factors such as comorbid pathology, the age range of patients in each particular registry, the level of maintenance of physical activity of patients after diagnosis, rehabilitation measures, and other factors. Thus, the mean age of patients in our registry was 37.8 years (range: 0-97 years). This value was lower than in registries in other countries: 47.82 years (range: 6-83 years) in the UK registry and 51 years (range: 39-62 years) in the Dutch registry.

The data collected through the self-reported questionnaire provide valuable insights into the course of the disease and the potential influence of various factors on its progression. The prevalence of comorbidities identified in our study (34.5%) was significantly higher than the rate reported in a previous study of the French registry (approximately 19.6%) and exceeded rates observed in other studies as well [40–42].

In our cohort, only one-third of patients reported integrating regular physical activity into their lifestyle, which is lower compared to findings from a large European survey of 1,147 respondents across 26 countries, where a greater proportion of patients reported using physiotherapy or occupational therapy (49%), as well as performing physical exercises (47%) [43].

According to the physical subscale score of the FDI, facial muscle function was generally preserved. This is consistent with the data on the onset of weakness: only 14.8% of patients who underwent a clinical examination at the center initially self-reported the onset of weakness in the facial muscles. However, after a detailed medical history, this proportion increased to 31.6%. Thus, the presence of facial weakness does not lead to significant functional limitations for patients and is not typically a reason to seek medical evaluation. Facial muscle weakness is often perceived as an individual characteristic or a variant of normal. The average GNEM-FAS score was 79.27, indicating generally relatively preserved functional abilities and quality of daily life. However, the range of scores varied from 19 to 100, demonstrating heterogeneity in the group, from severe cases to patients without functional limitations. The average score for the "Mobility" subscale was 71.7% of the maximum value, indicating reduced mobility in the patient cohort. The average scores for the "Upper Extremities" and "Self-Care" subscales also indicate preserved upper extremity function and self-care. However, the observed range of scores across all three scales also demonstrates that patients with FSHD have different domains of quality of life affected, suggesting heterogeneity in the sample affected. Efforts are currently underway to establish an online platform for self-reported questionnaires and to revise the existing scales, aimed at enhancing data collection and enabling more effective monitoring of disease progression in the registry.

Overall, the results obtained using the clinical assessment form are consistent with well-known and previously published clinical features of FSHD [44]. The most frequent clinical signs in the Russian registry were scapular winging, facial weakness, asymmetrical involvement, pectoral muscles atrophy.

Among the rare signs such as joint contractures, retraction of the Achilles tendons, dysarthria, ptosis and camptocormia. The group of patients with camptocormia was predominantly composed of older patients and had distinctive characteristics from the group of previously reported patients with camptocormia [45]. Our patients with camptocormia have an earlier age of onset, a smaller number D4Z4 RUs, and absence of neck muscle weakness in all cases. A previous study has described camptocormia as a distinct phenotypic variant of FSHD in elderly patients, based on the presence of specific clinical, genetic, and radiological features in this group. However, our findings suggest that this condition may be a rare, atypical symptom rather than a distinct disease phenotype. To confirm this hypothesis, a larger number of patients with FSHD-associated camptocormia and further study of their clinical characteristics are needed.

Correlation analysis between the number of D4Z4 RUs and clinical severity scales including age-corrected FSHD-CS and age-corrected CSS revealed moderate inverse correlation with high statistical significance. Overall, these results are consistent with previous correlation analyses in other studies, which also found a moderate inverse correlation between clinical severity scales and D4Z4 RUs [20, 27, 46].

Disease severity was significantly higher in probands than in their affected relatives with FSHD1. This predominance of more severe forms among probands likely reflects selective referral, as patients with pronounced symptoms are more frequently referred to medical attention and thus constitute the clinical cohort. Our overall data are consistent with a previously published study that also observed a higher mean FSHD-CS score in probands compared to relatives [47].

The distribution of clinical categories across RUs groups aligns with previously published data demonstrating that an increase in the number of repeat units correlates with decreased expressivity and penetrance [6, 7, 48]. The observation that category C (subjects without motor impairment or with minor possible FSHD-linked signs) was present exclusively in the 7–10 RUs group supports this relationship. Furthermore, the statistically significant differences between the 4–6 and 7–10 RU groups highlight a clinical threshold of approximately 7 RUs for incomplete penetrance. The presence of category D patients (atypical signs) across all three RUs groups may reflect both severe forms with early onset and the broad phenotypic spectrum of disease expression, potentially influenced by modifiers that warrant further investigation.

In a study of Banerji et al (2020), cluster analysis involving 222 patients was based on the age of onset of each of four muscle weakness symptoms (facial, periscapular shoulder, foot dorsiflexor, and hip girdle weakness [35]. In contrast, our cluster analysis employed delta scores as time points rather than absolute age of onset, allowing for a more dynamic assessment of disease progression. This approach identified three distinct patient groups with differing disease trajectories. The largest group corresponded to the classic phenotype, characterized by early onset and relatively rapid progression. The two smaller clusters exhibited slow progression but differed in their initial presentation: one began with facial weakness, while the other started with periscapular involvement and delayed facial symptoms. Identifying these distinct progression patterns has important clinical implications. Recognizing the existence of slowly progressive phenotypes may facilitate earlier diagnosis, particularly in patients who might otherwise be misdiagnosed or remain under observation for extended periods. Moreover, these findings contribute to a more comprehensive understanding of the natural history of FSHD, highlighting its clinical heterogeneity and broadening the spectrum of recognized disease variants.

## 5 Conclusion

Here we presented the first comprehensive description of the Russian FSHD Patient Registry with the cohort size comparable to that of many international registries. Overall, the clinical and genetic profile of our cohort is consistent with data reported from other countries. However, some notable differences were also observed. The Russian registry demonstrates a predominance of alleles with 3 RUs, and a younger median age of participants. The younger cohort age likely explains the less proportion of extramuscular findings and lower proportion of patients with ambulation loss.

The obtained data facilitate further studies and the expansion of the FSHD patient cohort. The combination of specialized clinical scales with self-administered questionnaires will enable more accurate longitudinal assessment of patients’ quality of life and further delineation of factors influencing disease prognosis.

## Statements and Declarations

### Ethics approval and consent to participate

This study was carried out in accordance with the Code of Ethics of the World Medical Association (the Declaration of Helsinki). The clinical and molecular genetic study was approved by the Institutional Review Board of the Research Centre for Medical Genetics, Moscow, Russia (Resolution No. 8/2, 2 November 2015).

### Consent to publish

Written informed consent to publish was obtained from the patient and from the patient’s parents.

### Data availability

All data produced in the present study are available upon reasonable request to the authors.

### Materials availability

Not applicable

### Code availability

Not applicable

### Author contribution

Conceptualization: Aysylu Murtazina, Mikhail Skoblov; Methodology: Aysylu Murtazina, Mikhail Skoblov, Darya Sherstyukova, Nikolay Zernov, Artem Borovikov; Data collection: Anna Kuchina, Aysylu Murtazina, Dmitrii Subbotin, Elena Dadali, Inna Sharkova, Galina Rudenskaya; Formal analysis and investigation: Anna Kuchina, Darya Sherstyukova, Artem Borovikov, Margarita Soloshenko; Writing - original draft preparation: Anna Kuchina, Darya Sherstyukova; Writing - review and editing: Aysylu Murtazina, Anna Kuchina, Artem Borovikov; Funding acquisition: Sergey Kutsev; Supervision: Aysylu Murtazina, Mikhail Skoblov, Sergey Kutsev

## Acknowledgments

We thank the physicians who referred patients for DNA diagnostics and contributed to the establishment of the Russian FSHD Patient Registry:

Olga Zinovieva (Moscow) (deceased)

Sergey Nikitin (Moscow)

Sergei Kurbatov (Voronezh)

Sergey Bardakov (Saint Petersburg)

Dmitry Druzhinin (Moscow)

Olga Gilvanova (Moscow)

Eugenia Melnik (Moscow)

Anait Voskanyan (Moscow)

Elena Shabanova (Saint Petersburg)

Nina Demina (Moscow)

Natalia Semenova (Moscow)

Evgenia Druzhinina (Moscow)

Elizaveta Petelina (Tomsk)

Ludmila Bessonova (Moscow)

Irina Mishina (Moscow)

Fatima Bostanova (Moscow)

Marina Petukhova (Moscow)

Natalia Kuryakova (Moscow)

