## Supplementary for "The Russian FSHD registry: a first look at the cohort"

**Supplementary data**

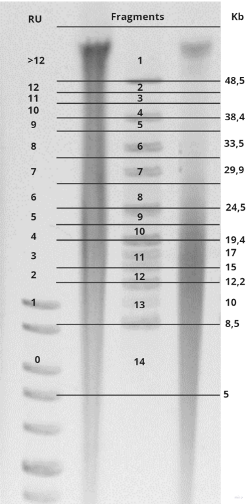

**Fig. S1** Gel image obtained after pulse-field electrophoresis (PFGE) and staining with ethidium bromide, captured using the “ChemiDOC XRS+ system” (Bio-Rad Laboratories, Veenendaal, The Netherlands). The schematic of gel fragmentation relative to the fragment lengths of the molecular weight markers—the Lambda Mix Marker (19) and the M12 DNA Ladder. The gel fragmentation scheme was designed so that each gel piece contains a D4Z4 array with a specific number of repeats

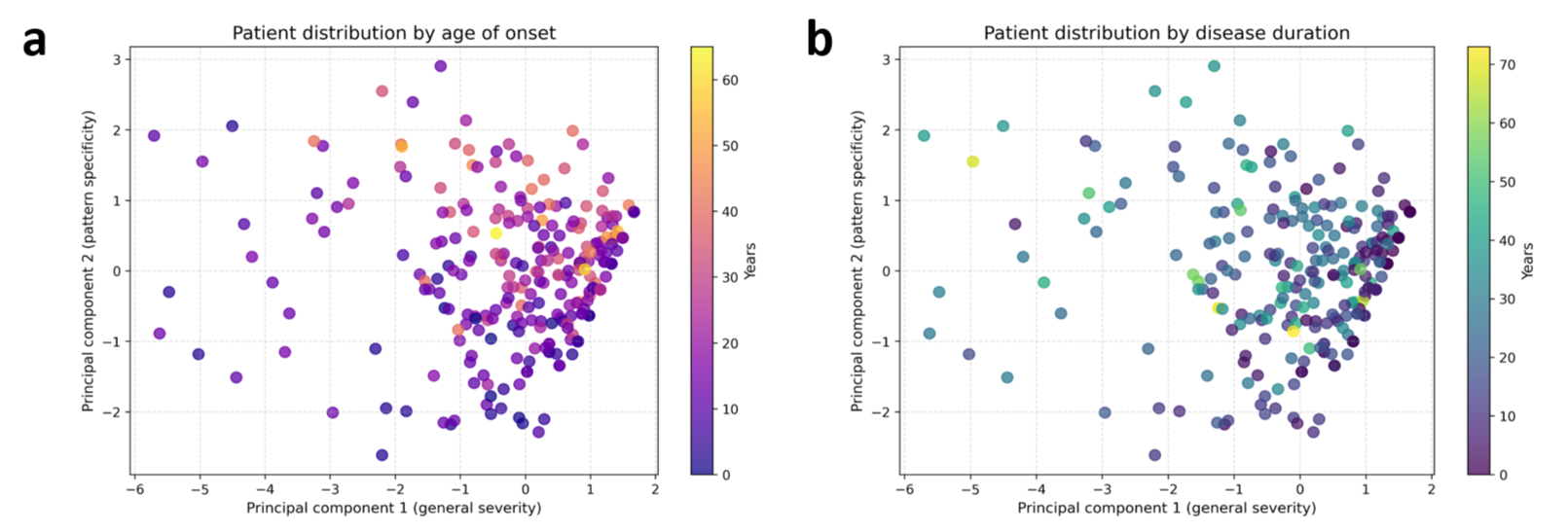

**Fig. S2** PCA-based visualization of patient distribution by clinical characteristics. (a) Patients are coloured by age of symptom onset (years). (b) Patients are coloured by disease duration (years). Two-dimensional principal component analysis (PCA) was applied to standardized manual muscle testing (MMT) scores across different muscle groups to visualize inter-patient variation in weakness distribution patterns. Each point represents one patient projected onto the first two principal components, which captured the largest sources of variance in the MMT data. MMT scores for muscle groups were developed as summarising of Medical Research Council (MRC) scores for the following muscles, which were examined bilaterally (right and left): (1) facial MMT: orbicularis oculi, orbicularis oris and cheek; (2) shoulder girdle MMT: biceps and triceps; (3) pelvic girdle and thigh MMT: hip flexors, quadriceps and hamstrings; (4) lower leg MMT: tibialis anterior, triceps surae and extensor hallucis brevis.

**Supplementary table 1** Clinical assessment form

| Clinical assessment form | |
| --- | --- |
| (I) Basic patient information | 1) Date of birth; 2) Gender; 3) Ethnicity; 4) Birth region; 5) Region of residence; 6) Age at examination; 7) Date of visit; 8) Family history. |
| (II) Genetic diagnostic results | 1) Diagnostic confirmation status; 2) The laboratory; 3) FSHD type; 4) The D4Z4 RUs number; 5) Pathogenic/likely pathogenic variant in the *SMCHD1* gene. |
| (III) Anamnestic data | 1) Age of disease onset; 2) First symptom of the disease; 3) Age of onset of the following symptoms: weakness of the facial muscles, shoulder girdle muscles, trunk muscles, pelvic girdle and hip muscles, and extensor muscles of the feet 4) Typical descending or ascending course of the disease. |
| (V) Neurological examination | 1) Asymmetry of involvement; 2) Muscle atrophy (supraspinatus and infraspinatus, trapezius, pectoralis, biceps, triceps, forearms, hands, paravertebral muscles, gluteals muscles, hamstrings, peroneals, gastrocnemius, foot muscles on both sides) and muscle hypertrophy in the legs; 3) Tendon reflexes assessment (biceps, triceps, brachioradialis, patellar, and ankle); 4) Sensory disturbances; 5) Cranial nerve function: oculomotor and bulbar impairments, such as ptosis, ophthalmoparesis, dysarthria, and dysphagia; 6) Musculoskeletal disorders (unilateral or bilateral scapular winging, lumbar hyperlordosis, thoracic deformities, joint contractures, Achilles tendon retractions, and foot deformities); 7) Axial and trunk involvement: dropped head, camptocormia, Beevor's sign. 8) Gait abnormalities (steppage gait, waddling gait, and performance on the heel-to-toe walking test); 9) Cognitive impairment; 10) Additional signs: Gowers' sign, grip myotonia and hand tremor. |
| (IV) Specialized clinical scales | 1) Сlinical severity scale (CSS) developed by Ricci et al. [[17]](https://www.zotero.org/google-docs/?4vxb9b) and age-corrected CSS [[18]](https://www.zotero.org/google-docs/?0AIPtj); 2) FSHD clinical score (FSHD-CS) and age-corrected FSHD-CS [[16,21]](https://www.zotero.org/google-docs/?FzblMQ); 3) Clinical Categories assessment [[19,20]](https://www.zotero.org/google-docs/?2D4GlT). 4) Muscle strength was assessed bilaterally using the Medical Research Council (MRC) scale (0–5) across 48 specific muscles. The evaluation encompassed facial muscles (orbicularis oculi and oris, buccinator), shoulder girdle and proximal upper limb muscles (supraspinatus, infraspinatus, deltoid, pectoralis, biceps, triceps), hand and forearm muscles (flexors and extensors of the wrist and fingers, interossei), pelvic and thigh muscles (hip flexors, extensors, adductors, and abductors; quadriceps; hamstrings), and distal lower limb muscles (peroneals, soleus, toe extensors, and extensor hallucis longus); 5) Total MMT score for 48 specific muscles, total MMT score for facial muscles, total MMT score for neck muscles,total MMT score for shoulder muscles, total MMT scores for pelvic and thigh muscles, total MMT scores for peroneal muscles. |
| (VI) Extramuscular signs | 1) Retinal angiopathy; 2) Cataracts; 3) Hearing loss; 4) Respiratory disturbances; 5) Others. |
| (VII) Laboratory and instrumental studies | 1) CK, liver enzymes level; 2) Electroneuromyography; 3) Electrocardiography; 4) Echocardiography 5) Brain MRI; 6) Muscle MRI; 7) Respiratory function tests. |

**Supplementary table 2** Clinical characteristics of patients with FSHD

| **Clinical sign** | **Count of patients, n** | **Percent, %** |
| --- | --- | --- |
| Asymmetrical involvement | 262 | 84.5 |
| **Atrophy muscles** | | |
| supraspinatus, infraspinatus, trapezius | 242 /310 | 78.1 |
| deltoid | 22 /310 | 7.1 |
| biceps | 127 /310 | 40.9 |
| triceps | 118 /310 | 38.1 |
| distal arm | 24 /310 | 7.7 |
| pectoral | 245 /310 | 79 |
| paravertebral | 178 /310 | 57.4 |
| gluteus maximus | 67 /310 | 21.6 |
| quadriceps | 115 /310 | 37.1 |
| hamstrings | 113 /310 | 36.5 |
| peroneal | 157 /310 | 50.6 |
| foot | 40 /310 | 12.9 |
| **Muscle weakness** | | |
| orbicularis oculi | 243 /310 | 78.4 |
| orbicularis oris and cheek | 283 /310 | 91.3 |
| neck flexors | 32 /310 | 10.3 |
| neck extensors | 7 /310 | 2.3 |
| supraspinatus | 176 /310 | 56.8 |
| infraspinatus | 179 /310 | 57.7 |
| deltoid | 40 /310 | 12.9 |
| biceps | 112 /310 | 36.1 |
| triceps | 68 /310 | 21.9 |
| wrist flexors | 15 /310 | 4.8 |
| wrist extensors | 36 /310 | 11.6 |
| hand finger flexors | 12 /310 | 3.9 |
| hand finger extensors | 46 /310 | 14.8 |
| interosseous | 38 /310 | 12.3 |
| hip extensors | 141 /310 | 45.5 |
| hip flexors | 121 /310 | 39.0 |
| hamstrings | 119 /310 | 38.4 |
| quadriceps | 71 /310 | 22.9 |
| adductors | 70 /310 | 22.6 |
| abductors | 49 /310 | 15.8 |
| tibialis anterior | 159 /310 | 51.3 |
| triceps surae | 59 /310 | 19.0 |
| extensor hallucis brevis | 143 /310 | 46.1 |
| extensor digitorum brevis | 127 /310 | 41.0 |
| **Others signs** | | |
| ptosis | 7 /310 | 2.3 |
| ophthalmoparesis | 1 /310 | 0.3 |
| dysarthria | 16 /310 | 5.2 |
| dysphagia | 3 /310 | 0.9 |
| scapular winging | 292 /310 | 94.1 |
| lumbar hyperlordosis | 164 /310 | 52.9 |
| chest wall deformities | 146 /310 | 47.1 |
| Beevor’s sign | 212 /310 | 68.4 |
| сamptocormia | 5 /310 | 1.6 |
| joint contractures | 21 /310 | 6.7 |
| foot deformities | 32 /310 | 10.3 |
| achilles tendon retraction | 22 /310 | 7.1 |
| cognitive impairment | 2 /310 | 6.5 |
| **Laboratory and instrumental studies** | | |
| elevated CK levels | 177 /216 | 81.9 |
| electroneuromyography | 225 /310 | 72.6 |
